# Frequency Of pRenatal CAre viSiTs (FORCAST): study protocol to develop a core outcome set for prenatal care schedules

**DOI:** 10.1101/2023.04.17.23288705

**Authors:** Mark Turrentine, Buu-Hac Nguyen, Beth Choby, Susan Kendig, Tekoa L. King, Milton Kotelchuck, Tiffany A. Moore Simas, Sindhu K. Srinivas, Christopher M. Zahn, Alex Friedman Peahl

**Affiliations:** Department of Obstetrics & Gynecology, Baylor College of Medicine, 6651 Main Street, Ste. F1020, Houston, TX 77030 USA; University of Michigan College of Literature, Science, and the Arts, 500 S. State St., Ann Arbor, MI 48109 USA; Department of Medical Education, Baptist Health Sciences University 1003 Monroe Ave. Memphis, TN 38104 USA; SSM Health-St. Louis Region, Maternal Services, 1191 Fortune Blvd., Shiloh, Il. 62269, USA; Department of Family Health Care Nursing, University of California, San Francisco School of Nursing, 2 Koret Way, San Francisco, CA 94143 USA; Department of Pediatrics, Harvard Medical School, 125 Nashua St., Boston, MA 02114 USA; Department of Obstetrics & Gynecology, Pediatrics, Psychiatry, and Population & Quantitative Health Sciences, University of Massachusetts Chan Medical School/UMass Memorial Health, 119 Belmont St., Worcester, MA 01605 USA; Department of Obstetrics and Gynecology, Perelman School of Medicine, University of Pennsylvania, Philadelphia, PA 19104 USA; Clinical Practice and Health Equity and Quality, American College of Obstetricians and Gynecologists, 409 12^th^ Street SW, Washington, DC 20024 USA; Department of Obstetrics and Gynecology and Institute for Healthcare Policy and Innovation, University of Michigan, 2800 Plymouth Rd. Ann Arbor, MI 48109 USA

**Keywords:** Core outcome set, Delphi, prenatal care schedules, telemedicine

## Abstract

**Background:** Prenatal care, one of the most common preventive care services in the United States, endeavors to improve pregnancy outcomes through evidence-based screenings and interventions. Despite the prevalence of prenatal care and its importance to maternal and infant health, there are several debates about the best methods of prenatal care delivery, including the most appropriate schedule frequency and content of prenatal visits. Current U.S. national guidelines recommend that low-risk individuals receive a standard schedule of 12 to 14 in-office visits, a care delivery model that has remained unchanged for almost a century.

**Objectives:** In early 2020, to mitigate individuals’ exposure to the SARS-CoV-2 virus, prenatal care providers implemented new paradigms that altered the schedule frequency, interval, and modality (e.g., telemedicine) of how prenatal care services were offered. In this manuscript, we describe development of a core outcome set (COS) that can be used to evaluate the effect of the frequency of prenatal care schedules on maternal and infant outcomes.

**Methods:** We will systematically review the literature to identify previously reported outcomes important to individuals who receive prenatal care and the people who care for them. Stakeholders with expertise in prenatal care delivery (i.e., patients/family members, healthcare providers, and public health professionals and policymakers) will rate the importance of identified outcomes in an online survey using a three-round Delphi process. A virtual consensus meeting will be held for a group of stakeholder representatives to discuss and vote on the outcomes to include in the final COS.

**Results:** The Delphi survey was initiated in July 2022 with 71 stakeholders invited. A virtual consensus conference was conducted on October 11, 2022. Data is currently under analysis.

**Conclusions:** More research about the optimal schedule frequency and modality for prenatal care delivery is needed. Standardizing outcomes that are measured and reported in evaluations of the recommended prenatal care schedules will assist evidence synthesis and results reported in systematic reviews and meta-analyses. Overall, this COS will expand the consistency and patient-centeredness of reported outcomes for various prenatal care delivery schedules and modalities, hopefully improving the overall efficacy of recommended care delivery for pregnant people and their families.

**Trial Registration:** This study was registered in the Core Outcome Measures for Effectiveness (COMET) database on January 18, 2022, registration #2021 http://www.comet-initiative.org/Studies/Details/2021.

## Introduction

Prenatal care, one of the most common preventive care services in the United States, endeavors to improve pregnancy outcomes through evidence-based screenings and interventions [1]. Despite the prevalence of prenatal care and its importance to maternal and infant health, there are several debates about the best methods of prenatal care delivery, including the most appropriate schedule frequency and content of prenatal visits [2]. Current U.S. national guidelines recommend that low-risk individuals receive a standard 12 to 14 in-office visit schedule, a care delivery model that has remained unchanged for almost a century. Other guidelines from peer countries, as well as national consensus panels, have recommended a reduced visit schedule based on recommended prenatal care services for average-risk people [3-5].

In early 2020, to mitigate individuals’ exposure to the SARS-CoV-2 virus, prenatal care providers implemented new paradigms that altered the schedule frequency, interval, and modality (e.g., telemedicine) of how prenatal care services were offered. COVID-19 pandemic-related changes to how prenatal care was provided accelerated existing plans of the American College of Obstetricians and Gynecologists (ACOG) to redesign prenatal care for low-risk pregnant people. ACOG urgently convened a multidisciplinary panel to review how prenatal care was administered to the average-risk individual [5]. The panel provided recommendations for: 1) prenatal visit schedules (care initiation, visit timing and frequency); 2) integration of telemedicine (virtual visits and home devices); and 3) care individualization. However, it was recognized that there were significant gaps in evidence needed to guide national policy change on prenatal visit schedules, including visit frequency and modality. Meta-analyses of studies that addressed the prenatal care visit schedules and the effects on maternal or infant outcomes have noted inconsistencies in which outcomes are routinely collected and reported [6-9]. There is a need for consistent reporting of maternal and infant outcomes in future clinical trials that compare the traditional 12 to 14 in-office visit approach with new models of prenatal care schedules that incorporate different frequency of visits and use of telemedicine.

Core outcome sets (COS) are key tools for ensuring consistent, homogenous reporting of outcomes across studies. Use of COS can result in improved clinical practice via standardization of outcomes across studies, thereby making it easier to compare outcomes. Furthermore, use of COS can reduce the risk of outcome-reporting preference, ensuring that all trials contribute functional information [10]. The objective of this study is to develop a COS to standardize outcome selection, collection, and reporting for future studies that compare prenatal care schedules with different frequencies of prenatal visits delivered across any modality (e.g., in-office care, group prenatal care, or telemedicine).

## Methods

### Steering Committee

This study has been prospectively registered with the Core Outcome Measures in Effectiveness Trials (COMET) initiative website, registration number 2021. We will follow reporting guidelines for developing core outcome sets, as outlined by the Core Outcome Set-STAndards for Development (COS-STAD) and follow the Core Outcome Set-STAndardised Protocol Items statement (COS-STAP) [11, 12]. A steering group including prenatal care providers, clinical researchers, and obstetric healthcare policymakers has been formed to guide the development of this COS. Members of the steering group were selected to represent various disciplines, geographical areas, and expertise. Within the steering committee, a study advisory board has been established, consisting of a study coordinator (BN) and two members of the steering committee (MT and AP) who will conduct the day-to-day management of the study.

### Definitions

The steering committee recommended that the COS should apply to clinical studies evaluating the frequency of prenatal care schedules for average-risk individuals receiving prenatal care when compared to current U.S. national guideline recommendations of a 12 to 14 in-office visit schedule. Average-risk was defined broadly using a prior consensus committee definition and includes all pregnant individuals seen by maternity care providers (e.g., obstetrician gynecologists, family medicine physicians, nurse practitioners, certified nurse-midwives), without significant comorbidities requiring exclusive care by a maternal fetal medicine physician [5]. The potential intervention(s) covered by the COS could include the recommended frequency of prenatal care visits in several modalities: 1) traditional 12 to 14 in-office appointments versus reduced number of in-office visits; 2) method of delivery of the frequency of various types of prenatal care visit recommendations (traditional 12 to 14 in-office visits versus a reduced number of visits provided by a hybrid of in-office combined with telemedicine modalities); and potentially 3) other innovative models of prenatal care schedule recommendations (traditional 12 to 14 in-office visits compared to a reduced number of scheduled visits using care delivery models such as group prenatal care or pregnancy medical homes that incorporate either in-office only or a hybrid of in-office and telemedicine.

### Study Overview

This study will be divided into three separate phases: 1) ascertaining potential core outcomes; 2) defining core outcomes for inclusion; and 3) consensus meeting.

#### Phase One: Developing a Preliminary Core Measurement Outcome Set

Two systematic reviews were recently commissioned for evaluating the effect of varying the frequency of prenatal care visit schedule recommendations on maternal and infant outcomes [5, 9]. To identify any additional outcome measures published in the literature, we will supplement these systematic reviews with an electronic investigation with no date or language boundary using the databases of MEDLINE via the PubMed interface, Web of Science, Embase (Excerpta Medica Database), and CENTRAL (Cochrane Central Register of Controlled Trials) to identify outcome measures in systematic reviews that reported on the effect of different frequency of prenatal visit recommendations on maternal and infant outcomes. We will evaluate unpublished collected works (i.e., “gray literature”) by searching Google Scholar and screening the first 100 results as is commonly done, based on the assumption that the most applicable results would emerge first.

We will also identify potentially relevant outcome measures from the medical literature to inform the consensus process. Data sources that may be considered are systematic reviews of published studies and reviews of published qualitative work that have evaluated the effect of the recommended frequency of scheduled prenatal visits on maternal and infant outcomes. A list of outcomes from published studies will be supplemented by a review of qualitative research studies investigating patients’ opinions regarding the frequency of prenatal care visits.

Steering group members will be provided with a preliminary outcome list, as well as a summary of the systematic review(s) and the strength of evidence for each outcome (based on either the Cochrane Handbook for Systematic Reviews of Interventions [13] or the Grading of Recommendations Assessment, Development and Evaluation (GRADE) [14]). The list of core outcomes will be piloted and adjusted by the steering group, incorporating both professional expertise and public member perspectives, before finalizing. The steering group members will be asked to “From the following list choose the ‘minimum standard outcomes’ that you think are important when developing a core outcome set for the Frequency Of pRenatal CAre viSiTs (FORCAST) recommended schedule.” Each respondent will be given the option to include, exclude, or combine each outcome with other outcomes that have similar clinical definitions (e.g., combining gestational age at birth and preterm birth into a single outcome) to limit the number of COS outcomes and increase the likelihood that the COS is applied in future research. Steering group members will also be given the opportunity to add outcomes they believe are critical for the COS. Steering group members will be encouraged to base their COS decisions on the strength of evidence reported in the medical and public health literature. Steering group member responses will be tabulated into an outcome inventory spreadsheet. Responses to the survey will be reviewed independently by the advisory board members, duplicate suggestions will be removed, and the remaining responses will be grouped into domains of Maternal or Infant outcomes. Additional outcomes proposed by the steering committee will be added to the COS if at least two members write in the outcome, per specifications of the COMET Handbook [10].

The steering committee will then review the domains and finalize a comprehensive list of outcomes. Following the steering group’s consensus, the list of outcomes will then be prepared for the Delphi survey [10]. All language for the COS will be developed using definitions from professional organizations and patient-facing resources tested across diverse populations where possible. Definitions will be piloted with three public members to confirm readability and comprehension.

#### Phase Two: Conducting a Delphi Survey to Gain Consensus Opinion on Items to Include in a Standardized COS

We will conduct a Delphi panel to develop the stakeholder-informed COS (10). In a COS structure, this method is used for attaining convergence of opinion from stakeholders on the importance of different outcomes in sequential questionnaires sent electronically. We will utilize sequential online questionnaires with participant feedback between rounds to incorporate the perspectives of diverse stakeholders who contribute to and receive prenatal care, including: 1) public members: persons who are currently using prenatal care, have utilized prenatal care in the past year, or provide non-medical support to pregnant and postpartum people (e.g., pregnant and postpartum people, family members); 2) prenatal care providers and researchers: healthcare providers who administer prenatal care (e.g., obstetrician/gynecologists, family medicine physicians, neonatologists, advanced practice practitioners, nurses) with representation from both academic and community health settings and investigators involved in perinatal research (e.g., clinical researchers, health services researchers); 3) public policy members: individuals involved in development of guidelines, regulation, and payment for pregnancy and postpartum care (e.g., clinical guideline developers, public health representatives, medical insurance payer representatives).

##### Recruitment

Potential participants will be invited to participate in the Delphi survey with a personalized email describing the project. Baseline demographic information (age, gender, race, ethnicity) and baseline practice information (if a prenatal care provider) will be obtained for each participant at the beginning of the survey. Participants will be asked to complete the survey in seven days, with a reminder email sent at the end of 14 days to prompt completion. Participation will be optional and consent to participate will be implied if a contributor responds to the survey. We aim to recruit a minimum of 20 public members, 20 providers and researchers (including a mixture of academic and community providers), and 20 public health and public policy members, with balanced representation across identities, including race and U.S. geographic regions.

##### Delphi Procedures

Before starting the survey, participants will be assigned a unique identifier to anonymize their responses and allow identification and linkage of individual responses in rounds of the Delphi exercise and feedback. Participants will be invited to provide their name and consent to be acknowledged as a member of the Delphi panel in the publication arising from this research. We anticipate a three-round survey; however, the round two results will be reviewed by the steering committee to consider the need for a further Delphi survey round. All participants who complete the first round of the Delphi exercise will be invited to participate in the second.

All participants who complete the second round will be invited to participate in the third. The first-round survey will include scoring of core outcome measurement items informed by the preliminary list from phase one. Outcomes will be ranked on a 1-9 Likert scale ranging from least important to critical importance (1-3 is of limited importance, 4-6 is important but not critical, and 7-9 is critical) [14]. In round 1, participants can suggest new items to be included in the second round. If two or more participants suggest an additional core outcome measure, it will be reviewed by the steering group and unique outcomes will be entered into round 2. All core outcome measures from round 1 will be carried forward to round 2. Participants will also be given the opportunity to share explanations for their ratings through online message boards.

Following round 1, the study advisory board will calculate the median and range of scores for each round 1 outcome by the stakeholder group, as well as the pooled median and range for all participants. Explanations shared through message boards will also be collated and shared with scoring summaries. For each round, the number of participants who were invited but do not participate (i.e., attrition rate) will be calculated and shown in the final publication of the COS. Prior to completing round 2, each participant will receive their own score on each potential core outcome measure, as well as the scores by stakeholder group and overall. Participants will be asked to reflect on their own scores and on the scores of other participants before re-scoring each individual outcome. After the round 2 survey has closed, the percentage of participants scoring each outcome will be calculated and tabulated for each individual, the stakeholder groups, and the overall pooled group. If 70% of the core outcomes have reached a consensus to be either included and/or excluded (as defined below) after round 2 of the Delphi survey, the third round of the Delphi will be omitted, and the group will proceed to the consensus meeting. If participants have not come to consensus at the conclusion of round 2, similar procedures will be repeated for round 3.

Consensus for inclusion will be defined as when ≥70% of participants scored the outcome “critical” for decision-making (a score of 7-9) and <15% of participants in the stakeholder group scored the outcome of “limited importance for decision-making” (a score of 1-3). Consensus for an outcome to be excluded from the COS will be defined when ≥70% of participants in the stakeholder group scored the outcome of “limited importance for decision-making” (a score of 1-3) and <15% of participants in the stakeholder group scored the outcome as “critical for decision-making” (a score of 7-9) [10]. Though there is no consensus regarding the ideal number of outcomes to include in a COS, based on existing literature and expert opinion, the steering committee has established a preset goal of 10 outcomes for the COS to enable implementation and use of the COS [15]. Items that reach consensus to be included (as defined above) for all three stakeholder groups will be automatically included in the final set of FORCAST COS recommendations, even if there are more than 10 core outcomes. Items that reach consensus to be excluded (as defined above) by all three stakeholder groups will be removed. If after the final round of the Delphi survey, no core outcome measures are excluded, the steering committee will determine outcomes to be brought forward to the consensus meeting using at least one of the following three criteria: 1) the mean outcome measure is scored as critical (a score of 7-9) by ≥70% of all participants included in the Delphi survey; 2) the top 10 outcome measures from the three different stakeholder groups (public members, prenatal care providers and researchers, and public policy members); or 3) outcome measures considered critically important by ≥70% of one or more of the different stakeholder groups [15].

#### Phase Three: Holding a Consensus Meeting to Identify the Main Items to be Included in the COS

A virtual consensus meeting will be held to review outcomes without consensus, resolve remaining discrepancies, and approve a final COS for the frequency of scheduled prenatal care visits. The consensus meeting will include at least seven of the nine steering committee members and three representatives from each of the three stakeholder groups (nine members), selected to maximize diversity, as voting members. Additionally, any participant who completed all three rounds of the Delphi survey will be invited to observe and provide their perspective in discussions. The meeting will begin with an initial briefing on the purpose and scope of the meeting, including a summary of the results from the initial rounds of the Delphi, outcomes with consensus for inclusion and exclusion, and outcomes where consensus has yet to be reached. The panel will begin with outcomes that reached consensus from all stakeholder groups for inclusion or exclusion, to approve those results. The panel will next consider items that did not reach consensus. These items will be the focus of the meeting. All consensus meeting participants will be given an opportunity to discuss each item and share their perspective on why it should or should not be included in the final COS. Members of the steering group and the three representatives from each stakeholder group will receive a summary of the results and be asked to independently consider items remaining that did not reach consensus to be either automatically included or excluded. The steering committee and stakeholder representatives will then take a majority vote as to whether or not the core outcome measure should be included in the final COS. While the preset goal of the consensus meeting is to encompass no more than 10 outcomes, if it is determined that more than 10 outcomes are necessary, these additional outcomes will be included. A summary of this process and the voting results will be published with the final report. Those outcomes excluded will be listed in the final publication. The final COS will be sent to all participants who participated in the three Delphi rounds in order to give them the opportunity to comment on the final results. The steering committee will draft the FORCAST COS guideline for publication and dissemination.

### Ethics Approval

The study was determined exempt by the University of Michigan institutional review board, as it was deemed non-human subject research (HUM00217486).

## Results

The Delphi survey was initiated in July 2022 with 71 stakeholders invited. A virtual consensus conference was conducted on October 11, 2022. Data is currently under analysis.

## Discussion

Applying a COS for the frequency of recommended scheduled prenatal care visits and mode of visit interaction is needed to assist forthcoming clinical studies, systematic reviews, and evidence-based clinical guidance to decide which prenatal care approach has the strongest efficacy and clinical utility, and for whom. Many prenatal care interventions, including laboratory tests, vaccinations, and routine screenings, are supported by robust randomized controlled trial evidence with consistent outcomes demonstrated through meta-analyses. However, schedules and methods of prenatal care delivery recommendations overall have not yet been subjected to these rigorous standards of assessment.

Maternity care stakeholders, including healthcare providers, health systems leaders, and payers, have numerous methods of prenatal care delivery to choose from, including the traditional visit schedule (visits every four weeks until 28 weeks, every two weeks until 36 weeks, and weekly until delivery); reduced visit schedules (8 to 9 visits based on necessary prenatal services); telemedicine; group prenatal care; and others. Establishing this COS may potentially generate a set of measures for assessing the relationship, effect, and magnitude of prenatal care visit schedules on maternal and infant health outcomes and help stakeholders to determine the comparative value of recommended prenatal care visit schedules. Establishing a COS is paramount to ensure pregnant individuals have access to the most effective prenatal care recommendations, delivered through the most efficient care delivery model.

## Data Availability

All datasets used and/or analyzed during the FORCAST study will be held by the FORCAST team at.

## Acknowledgements

We wish to thank Lamiya Ahmed and Megan McReynolds from the American College of Obstetricians and Gynecologists for organizational support of this project and providing lay person feedback for core outcome set definitions as well as editing of the manuscript as public member representatives. We also wish to thank Sarah Block for her assistance with the preparation of this manuscript.

## Data Availability

All datasets used and/or analyzed during the FORCAST study will be held by the FORCAST team at.

## Disclaimer

None

## Authors’ Contributions

MT and AFP are the Chief Investigators; they conceived the study, led the proposal and protocol development. All authors (MT, BN, BC, SK, TK, MK, TMS, SS, CZ, AFP) made substantial contributions to the conception and design of the Core Outcome Set study. BN was the lead trial methodologist. MT and AFP drafted the manuscript. All authors commented on revisions to draft versions of the manuscript, read, commented on, and approved the final manuscript. All authors are members of the overall FORCAST study team.

## Abbreviations

FORCAST: Frequency Of pRenatal CAre viSiTs
COS: core outcome set
COMET: Core Outcome Measures for Effectiveness
ACOG: American College of Obstetricians and Gynecologists

